# Changes in lipids and inflammation in adults with super-refractory status epilepticus on a ketogenic diet

**DOI:** 10.1101/2021.02.23.21252291

**Authors:** Alex M Dickens, Tory Johnson, Santosh Lamichhane, Anupama Kumar, Carlos A. Pardo, Erie G. Gutierrez, Norman Haughey, Mackenzie C. Cervenka

## Abstract

**Objective:** To test the hypothesis that increased ketone body production with a ketogenic diet (KD) will correlate with reductions in pro-inflammatory cytokines, lipid subspecies, and improved clinical outcomes in adults treated with an adjunctive ketogenic diet for super-refractory status epilepticus (SRSE).

**Methods:** Adults (18 years or older) were treated with a 4:1 (fat:carbohydrate and protein) ratio enteral KD as adjunctive therapy to pharmacologic seizure suppression for SRSE. Blood and urine samples and clinical measurements were collected at baseline (n=10), after 1 week (n=8), and after 2 weeks of KD (n=5). Urine acetoacetate, serum β-hydroxybutyrate, lipidomics, pro-inflammatory cytokines (IL-1β, IL-6), chemokines (CCL3, CCL4, CXCL13), and clinical measurements were obtained at these 3 time points. Univariate and multivariate data analyses were performed to determine the correlation between ketone body production and circulating lipids, inflammatory biomarkers, and clinical outcome.

**Results:** Changes in lipids included an increase in ceramides, mono-hexosyl ceramide, sphingomyelin, phosphocholine, and phosphoserines, and there was a significant reduction in pro-inflammatory mediators IL-6 and CXCL13 seen at 1 and 2 weeks of KD. Higher blood β-hydroxybutyrate levels at baseline correlated with better clinical outcome however, ketone body production did not correlate with other variables during treatment. Higher chemokine CCL3 levels following treatment correlated with greater length of intensive care unit stay, higher modified Rankin Scale score (worse neurologic disability) at discharge and 6-month follow up.

**Conclusions:** Adults receiving an adjunctive enteral ketogenic diet for super-refractory status epilepticus have alterations in select pro-inflammatory cytokines and lipid species that may predict response to treatment.

## Introduction

Status epilepticus (SE) is the second most frequent neurologic emergency worldwide^1-3^. Super-refractory status epilepticus (SRSE)^4^, SE that persists despite 24 hours of aggressive intervention, carries an in-hospital mortality rate above 40%^5, 6^. The ketogenic diet (KD) is a high fat, carbohydrate-restricted diet that shows promise for treating SRSE and may do so through fatty acid metabolism, ^7^,, ^8-12^, ^13, 14^. A study using a 4:1 ratio KD in 15 adults with SRSE found that 73% had resolution of SRSE, and more recent studies have replicated these findings^12, 15, 16^.

The exact mechanisms by which KD stops status epilepticus remain under investigation. During prolonged status epilepticus, systemic inflammation leads to loss of blood-brain-barrier integrity, leakage of neurotoxic serum proteins into the brain, and loss of inhibitory interneurons, all promoting further neuronal hyperexcitability^17-21^. Specific pro-inflammatory cytokines and chemokines including IL-1β, IL-6, CCL2, and CCL3 significantly increase with disease progression^22-32^. KD reduces inflammation thereby potentially halting this cyclical progression of neuronal injury and death^33-38^.

Another potential mechanism of action of KD may be lipid modulation that occurs when fatty acids are utilized during carbohydrate restriction. Certain lipid subspecies have been shown to be anti-inflammatory which may also explain the impact of KD on the inflammatory cascade^38^. This study examines the relationship between systemic inflammation, lipid modulation, and clinical outcomes in adults on a KD for management of SRSE. We hypothesized that increased ketone body production would correlate with reduction in pro-inflammatory cytokines, pro-inflammatory lipid subspecies, and improved clinical outcomes.

## Methods

### Population and demographics

The patients included in this study have previously been described^39^. Briefly, 15 patients with SRSE were recruited into a phase I/II clinical trial, and of these, 10 were included in this study. The ethical standards committee on human experimentation Johns Hopkins Medicine Institutional Review Board (JHM IRB) approved this experiment using human participants including biospecimen collection and participation in the clinical trial (JHM IRB study numbers NA_00003551 and <u>NA_00073063</u>, respectively). Written informed consent was obtained from all participants (or guardians of participants) in the study (consent for research). The original clinical trial from which participant biospecimens were collected was reported on ClinicalTrials.gov (NCT01796574). The patients were treated with a 4:1 (fat: carbohydrate and protein in grams) ratio KD upon diagnosis or hospital transfer. Clinical characteristics included participant age, gender, race, history of epilepsy prior to admission, and etiology of SRSE. “Ketosis” was defined as urine acetoacetate ≥ 40 mg/dL and/or serum β-hydroxybutyrate ≥ 2 mmol/L in order to calculate time to ketosis as a clinical outcome variable. Aliquots of plasma were collected from the patients repeatedly over time at 1-week intervals when possible. The plasma was stored at -80 °C prior to analysis. Due to the low number of samples longitudinally, we categorized the patients into 4 groups defined as follows:

- Baseline: Blood samples taken prior to the start of the ketogenic diet (n=10)
- Week 1: Blood samples taken after 1 week of ketogenic diet (n=8)
- Week 2: Blood samples taken after 2 weeks of ketogenic diet (n=5)

### Plasma lipid extraction

The lipids were extracted from the plasma using a modified Bligh Dyer approach. Throughout the procedure all steps were performed with glass to avoid extracting lipid like structures form lab plastic ware. All solvents used were of ultra-pure HPLC grade. 30 μL of plasma was mixed gently with 970 mL ddH_2_O and 2.9 mL mix of MeOH:DCM (2:0.9 v/v) to form a monophasic mixture. The organic fraction had a mix of the following lipids as internal standards: N-lauroyl-D-erythro-sphingosine (Cer d18:1/12:0, 6 ng/mL), 1,3(d5)-dihexadecanoyl-glycerol (d5-DAG d16:0/16:0, 12.5 ng/mL), D-galactosyl-β-1,1’ N-lauroyl-D-erythro-sphingosine (GlcCer d18:1/12:0, 3.3 ng/mL), D-lactosyl-β-1,1’ N-lauroyl-D-erythro-sphingosine (LacCer 18:1/12:0, 10.6 ng/mL), 1,3(d5)-dihexadecanoyl-2-octadecanoyl-glycerol (D-5 TAG 16:0/18:0/16:0, 0.5 ng/mL), cholesteryl-d7 palmitate (cholesteryl-d7 ester 16:0, 30 ng/mL), 1,2-dilauroyl-sn-glycero-3-phosphate (sodium salt) (PA d12:0/12:0, 1025 ng/mL), 1,2-dilauroyl-sn-glycero-3-phosphocholine (PC 12:0/12:0, 0.2 ng/mL), 1,2-dilauroyl-sn-glycero-3-phosphoethanolamine (PE d12:0/12:0, 1.6 ng/mL), 1,2-dilauroyl-sn-glycero-3-phospho-[1’-rac-glycerol] (PG d12:0/12:0, 200 ng/mL), 1,2-dilauroyl-sn-glycero-3-phospho-L-serine (PS d12:0/12:0), N-lauroyl-D-erythro-sphingosylphosphorylcholine (SM d18:1/12:0, 0.3 ng/mL), all internal standards were purchased from Avanti Polar Lipids, Inc. (Alabaster, AL). 1 mL of ddH_2_O and 0.9 mL of DCM was added to the mixture and the sample was briefly vortexed resulting in a biphasic mixture. This mixture was allowed to stand for 30 mins on ice prior to centrifugation (10 min, 3000g, 4 °C). The organic phase was then removed and stored at -20 °C until the mass spectrometry analysis.

### Mass spectrometry analysis

Prior to the mass spectrometry analysis 1 mL of the organic extract was evaporated to dryness under a stream of nitrogen and re-suspended in the running solvent (250 μL, MeOH:DCM, 1:1, containing 5 mM NH_4_CH_3_). The running solvent also contained 5mg/mL of ceramide C17:0 as a further internal standard to monitor instrument performance over time independently of the extraction process. The concentration of the lipids were then measured using an MS/MS^ALL^ experiment in positive mode on a TripleTOF™ 5600 (Sciex, USA) mass spectrometer. This is a data independent acquisition mode where the first quad steps through all masses from 200 to1200 Da in 1 Da steps. The selected precursor ion was then fragmented and quantified by the TOF with a scan range of 100-1500 Da. The accumulation time was set to 450 ms and the data was acquired using Analyst 1.7 TF (Sciex, USA) software. The mass calibration was performed after the run based on the internal standards within the sample. The sample (50 μL) was continually infused into the spectrometer at a constant flow rate (5 μL/min) using a LC-20AD pump and SIL-20AC XR HPLC system (Shimadzu, USA). The source parameters of the mass spectrometer was set up as follows: ion source gases 15 psi (GSI) and 20 psi (GS2), curtain gas 30 psi, temperature 150 °C, positive ion spray voltage +5500V, declustering potential at 80 V and collision energy at 10V. All samples were run in duplicate.

### Identification of lipids

In order to build a targeted method to extract out the identified lipids from each sample, a pooled sample was run eight times using the same mass spectrometry method. The lipids were then identified based on their fragmentation patterns using Lipid View (Sciex, USA). Each lipid species identified had to appear in 7 out of the 8 replicates and have a coefficient of variation below 20% in order for it to be included in the targeted lipid list. The resultant lipid list was then used to create a targeted method for extracting these specific lipids in Lipid View (Sciex, USA). The method was then applied to all the samples using MultiQuant v3.0 (Sciex, USA). The lipids intensities were corrected to their relevant class based internal standard. If the duplicate samples varied by more than 30% in intensity then the samples were re-run. For statistical analysis zero intensities were imputed by dividing the lowest intensity lipid by a factor of 1000.

### Cytokine and chemokine quantification

Serum IL-1β, IL-6, TNFα, CCL3, CCL4, and CXCL13 concentrations were measured serially in six participants by microbead multiplexed assays (Luminex®) per the manufacturer’s instructions. Only those participants that were not treated with concomitant immunosuppression were included in cytokine and chemokine analyses. Cytokine levels were measured prior to initiation of the ketogenic diet and at one and or two weeks after diet initiation when possible.

### Statistical analysis

The univariate analysis of lipid changes over time was assessed using a Wilcoxon rank test with multiple test correction. PLS-DA multivariate statistical analysis was performed using the PLS toolbox v 8.5 (Eigenvector labs, USA) for MATLAB 2018a (MathWorks, USA). The PLS-DA models were cross validated with a leave-one-out approach due to the low number of samples. Block variance scaling were used to scale the multiblock data. Multiblock-ANOVA-simultaneous component analysis (ASCA) a multivariate extension of ANOVA analysis was performed to allow interpretation of the variation induced categorical factors including Case (Baseline vs Week1), Sex, Race and history of epilepsy prior to admission. This multivariate analysis was also done using the PLS Toolbox 8.2.1 (Eigenvector Research Inc., Manson, WA, USA) in MATLAB 2017b (MathWorks, Inc., Natick, MA, USA).

All univariate analysis and resultant bar graphs were performed in Prism v 7.04 (GraphPad, USA). For the 4 group comparisons one way-ANOVA tests were used and multiple comparisons were corrected using the Benjamini and Hochberg FDR. Significant changes were defined as q values less than 0.05. Spearman correlation coefficients were calculated using the statistical toolbox in MATLAB 2017b and p-values < 0.05 (two-tailed) were considered significant for the correlations. The individual Spearman correlation coefficients (R) were illustrated circular correlation plot using the ‘corrplot’ package for the R statistical programming language.

For any missing values the half minima of that variable was used impute the value.

Cytokine and chemokine results are presented as index to control with the pre-ketogenic baseline measure set to 100%. Per previously described analyses strategies, if a measurement was below the limit of detection for a patient it was assigned the value of half the limit of detection and if more than one measurement was below the limit of detection for the same patient that patient was excluded from analysis ^40^. Measurements were assessed for statistical outliers and outliers (patient 8 CCL3 measurement, patient 1 CCL4 measurement, patient 10 TNF-α measurement, and patient 3 CXCL13 measurement) were not included in analyses but are presented in the figures. Decreases in cytokine or chemokine concentrations from baseline after initiation of the ketogenic diet were compared using a one-tailed paired t-test with a P < 0.05 considered statistically significant. Statistical analyses were performed with GraphPad Software version 8.3.1.

### Model-Based Clustering

Clustering of the lipidomic data was applied by using the *mclust* R package (version 5.4.5). *mclust* is a model-based clustering method where the model performances are evaluated by the Bayesian information criterion. Generally, the model with the highest Bayesian information criterion is chosen.

## Results

### Circulating lipids and cytokines

There were a total of 487 lipids detected and identified in serum from study participants. In addition, a panel of 6 cytokines and chemokines were also measured from the same samples. See Figure 1 for the overall experimental design.

**Figure 1.**
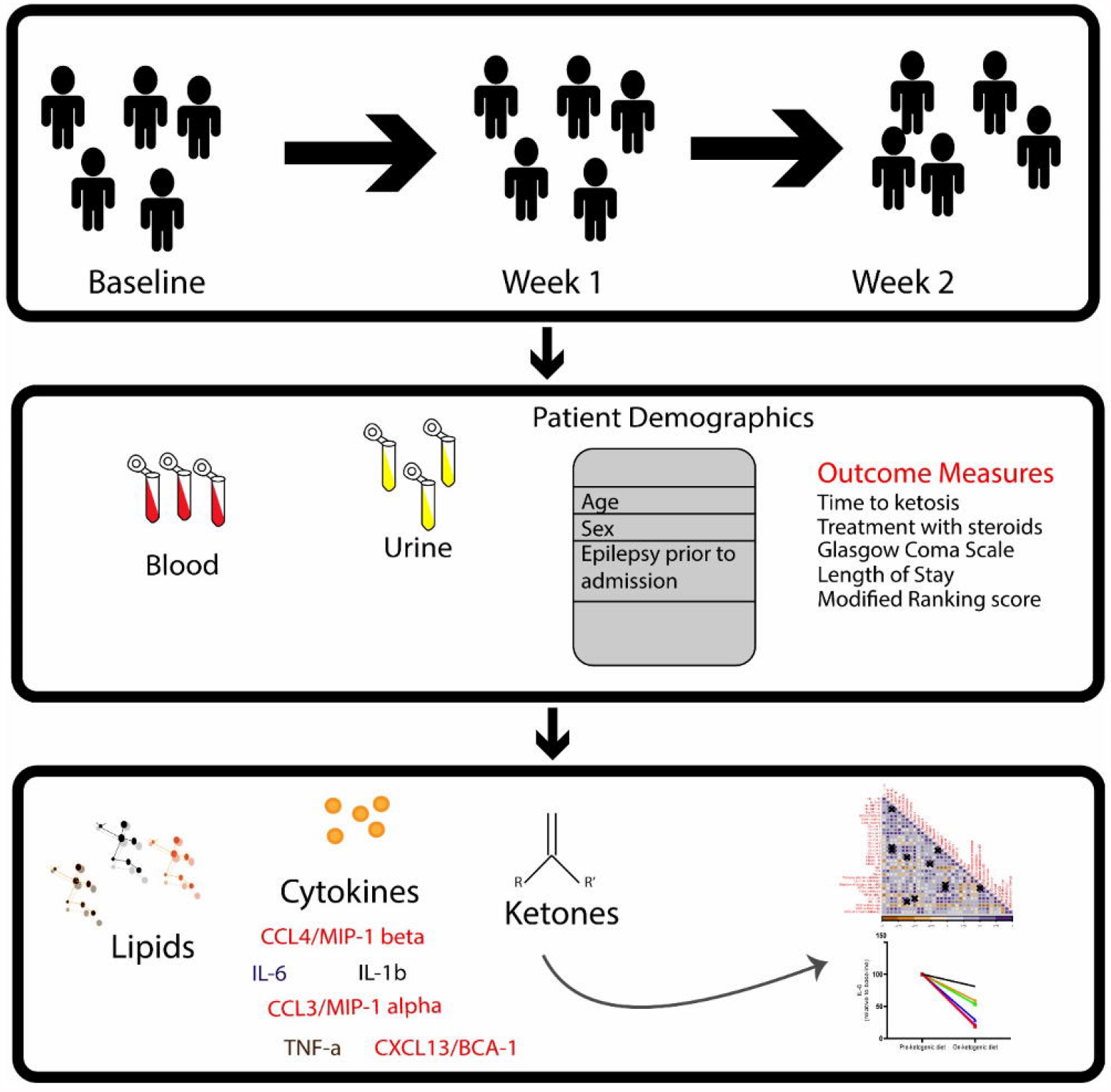
Study Design. In this study, patients with SRSE were recruited into a phase I/II clinical trial and treated with a 4:1 (fat:carbohydrate and protein in grams) ratio ketogenic diet (KD). Samples (blood and urine) and clinical measurements were collected at baseline: prior to KD (n=10), Week 1: after 1 week of KD (n=8), and Week 2: after 2 weeks of KD (n=5). Ketosis was confirmed by urine acetoacetate ≥ 40 mg/dL and/or serum β-hydroxybutyrate ≥ 2 mmol/L. Lipidomics, cytokine and chemokine measurements were performed in longitudinal settings. Then, univariate and multivariate data analyses were performed to determine the potential impact of diet on circulating lipids and inflammatory biomarkers.

### Lipid changes with ketogenic diet

Using a univariate approach, we identified a total of nine lipids that changed in abundance from baseline to week 1, and 29 lipids that changed in abundance by week 2, with q = 0.1 (see Table 1). Unknown triacylglycerol (TAG) species (54:11) and TAG (54:5) consistently increased from baseline to week 1, and from week 1 to week 2.

**Table 1.**

| Lipid | Week 1 Q | Week 2 Q |
| --- | --- | --- |
| Unknown TAG species (54:11) | 0.090 | 0.094 |
| TAG (58:7) | 0.090 | 0.236 |
| TAG (54:4) | 0.090 | 0.134 |
| TAG (54:5) | 0.090 | 0.070 |
| TAG (54:4) | 0.090 | 0.134 |
| TAG (54:3) | 0.090 | 0.276 |
| PS_O (34:6) | 0.090 | 0.386 |
| HexCer(30:0;2) | 0.090 | 0.457 |
| TAG (58:5) | 0.090 | 0.276 |
| TAG (54:4) | 0.110 | 0.078 |
| TAG (54:5) | 0.110 | 0.081 |
| TAG (56:4) | 0.110 | 0.081 |
| TAG (56:5) | 0.110 | 0.092 |
| TAG (56:7) | 0.116 | 0.022 |
| TAG (56:5) | 0.116 | 0.078 |
| HexCer (26:1;3) | 0.116 | 0.081 |
| Unknown TAG species (54:9) | 0.116 | 0.092 |
| Unknown TAG species (54:10) | 0.126 | 0.078 |
| TAG (56:6) | 0.186 | 0.022 |
| TAG (56:4) | 0.186 | 0.081 |
| TAG (56:7) | 0.199 | 0.015 |
| TAG (56:6) | 0.207 | 0.081 |
| Unknown TAG species (54:11) | 0.222 | 0.081 |
| Unknown TAG species (52:11) | 0.229 | 0.015 |
| TAG (56:5) | 0.250 | 0.078 |
| HexCer (40:0;3) | 0.265 | 0.022 |
| Unknown TAG species (50:10) | 0.277 | 0.034 |
| Cer (42:1;3) | 0.280 | 0.097 |
| PS (38:0) | 0.305 | 0.081 |
| TAG (56:5) | 0.308 | 0.078 |
| Unknown TAG species (52:10) | 0.310 | 0.078 |
| Unknown TAG species (54:4) | 0.322 | 0.078 |
| TAG (48:3) | 0.336 | 0.081 |
| TAG (52:6) | 0.353 | 0.081 |
| Unknown TAG species (52:10) | 0.393 | 0.078 |
| Unknown TAG species (52:10) | 0.396 | 0.081 |

Building an ASCA model using the individual lipidomic data from available samples we observed a clear drift in the multivariate models (Figure 2A), suggesting that there was a clear change in the serum lipid content after participants were placed onto a ketogenic diet that persisted over time. To further explore this change in serum lipid content over time we utilized an ANOVA-simultaneous component analysis (ASCA) multivariate model. This model can determine the effects of different independent variables such as time within the data in a manner similar to a standard two-way ANOVA. The ASCA modelling (Figure 2B) showed a distinct effect of time on ketogenic diet (effect = 12.5%) and gender (6.3%). Examination of the loadings of the first principal component for the factor of time on ketogenic diet demonstrated that several classes of lipids increased over time including ceramides, mono-hexosyl ceramides, sphingomyelins, phosphocholines, and phosphoserines (Figure 2C).

**Figure 2.**
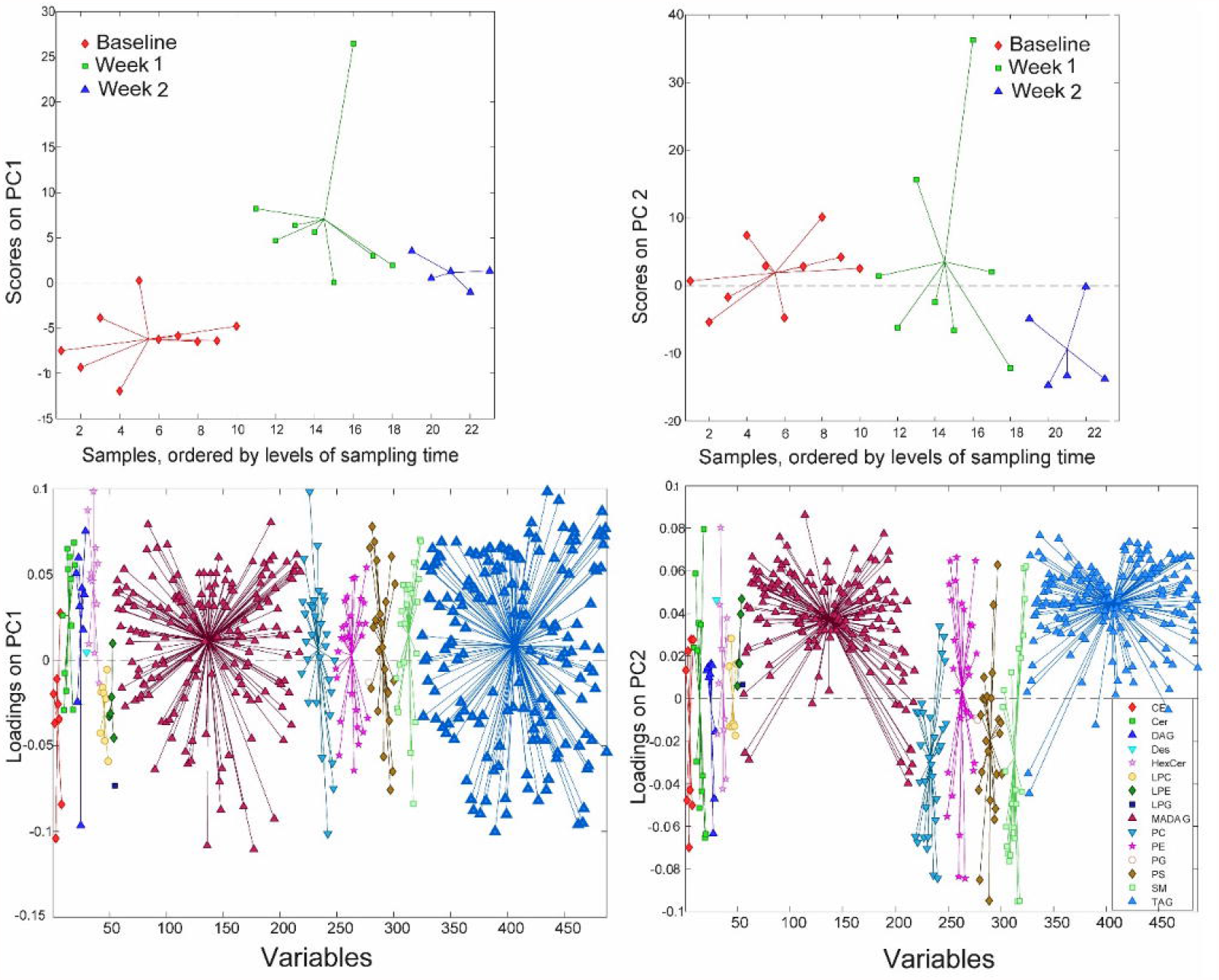
Overview of the lipidome in the longitudinal samples. A) ASCA score plots. B) PC1 score plot based on ASCA. These scores represent the lipidomics dataset arranged according to time of sampling in the ASCA score plot. Here each sample is represented by a point and colored according to the time of sample collection. Samples with similar scores are clustered together. (C) The corresponding PC1 loading plot. The loadings explain the pattern seen in the score plot which provides the means to interpret the class specific lipid alterations related to KD.

From the univariate model there was a clear change in the triglyceride species. Therefore, we plotted how these changed over time. Interestingly the short chain unsaturated lipids decreased in concentration whilst on the diet whereas the longer and more saturated compounds increased (Figure 3). These shorter chain saturated TAGs result from *de novo* lipogenesis suggesting that the ketogenic diet is actively supressing this metabolic pathway. The larger unsaturated species result from the diet and this is to be expected given the diet these patients are on.

**Figure 3.**
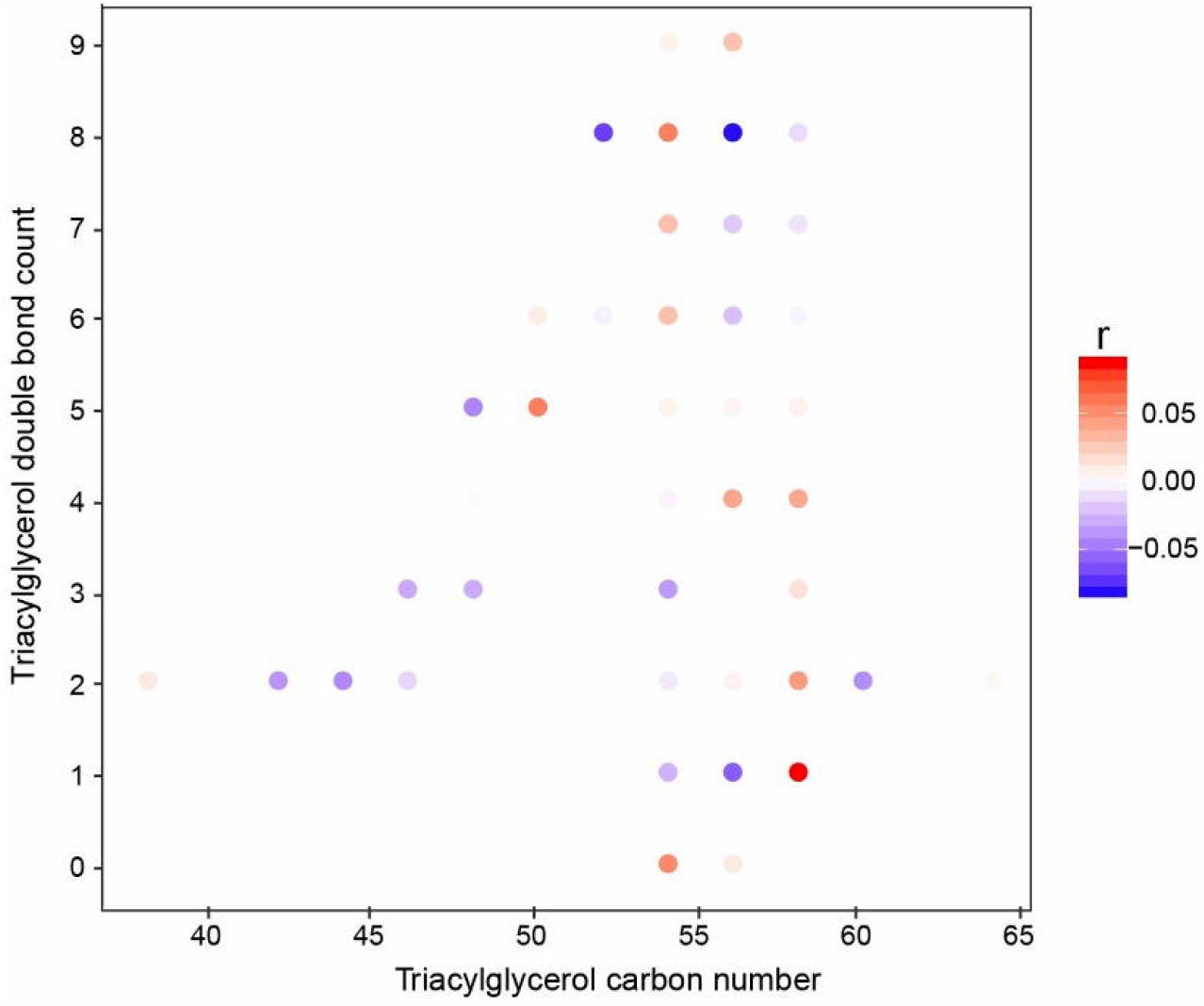
Correlation of individual TGs with the study week (baseline to week 2). The x-axis is the acyl carbon number and the y-axis is the acyl double-bond count. The dots are coloured according to the spearman coefficient.

### Serum cytokine changes over time

Two cytokines, IL-1β or TNFα were excluded from further analysis as only two participants had measurable levels above the limits of detection for the assay. IL-6 (n=5), CCL3 (n=5), CCL4 (n=6), and CXCL13 (n=6) concentrations were reliably measured in paired samples in participants and concentrations of each cytokine were normalized to individual participant at baseline and compared after one to two weeks on the ketogenic diet (Figure 4). Despite the small number of samples with detectable levels of cytokines pre- and post-treatment, there was a significant reduction in IL-6 (Figure 4A) at 1-2 weeks post-ketogenic diet compared to baseline (mean ± SD, pre-diet 100 ± 0 versus post-diet 43.8 ± 24.7, p=0.0013, one-way paired t-test). No differences in CCL3 (Figure 4B) (mean ± SD, pre-diet 100 ± 0 versus post-diet 166.1 ± 302.6, P=0.3458, one-way paired t-test) or CCL4 (Figure 4C) (mean ± SD, pre-diet 100 ± 0 versus post-diet 85.87 ± 22.02, P=0.1124, one-way paired t-test) were detected, although previously found to decrease in one preclinical studies of ketogenic diet^41^. An outlier (patient three) was detected in measurements for CXCL13 (Figure 4D) and therefore this patient was excluded from analyses. There was a significant reduction in CXCL13 after the ketogenic diet as compared to pre-diet (mean ± SD, pre-diet 100 ± 0 versus post-diet 58.36 ± 33.56, P=0.025, one-way paired t-test).

**Figure 4.**
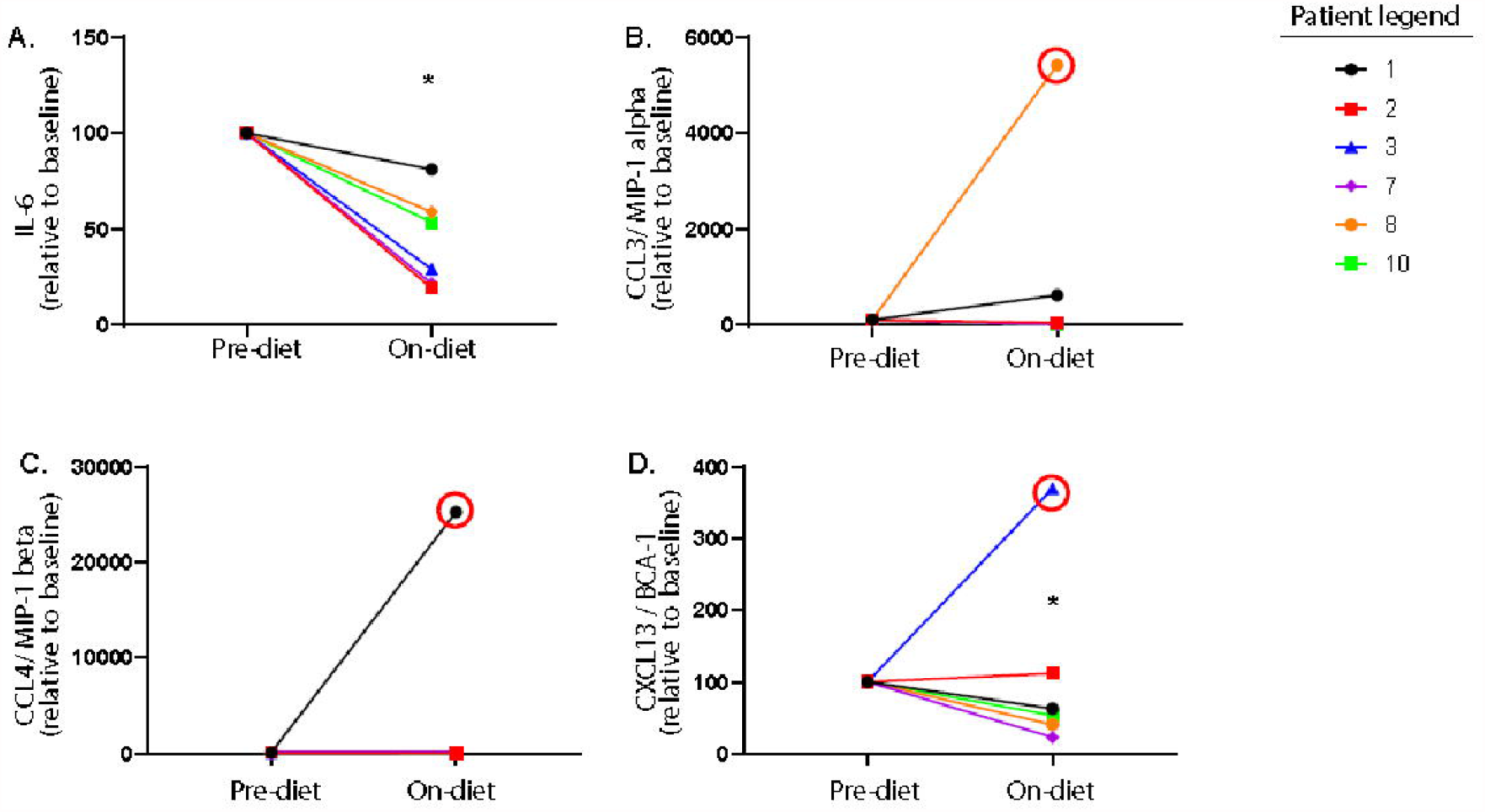
The ketogenic diet reduces pro-inflammatory cytokines in patients with refractory status epilepticus. Serum levels of (A) IL-6, (B) CCL3, (C) CCL4, and (D) CXCL13 were measured before patients initiated the ketogenic diet (pre-ketogenic diet) and one to two weeks after diet initiation (on-ketogenic diet). Data are expressed as the index of each cytokine with each patient baseline set to 100 (arbitrary units). Data were analyzed by a one-way paired t-test. There was a significant reduction in IL-6 (*P=0.0047) and CXCL13 (*P=0.025) in patients on the diet as compared to their baseline samples, but not in CCL3 (P=0.0854) or CCL4 (0.1124). Samples found to be statistical outliers are circled in red and are not included in analyses.

### Associations between serum lipids and cytokines

Due to the large number of lipids detected in the serum and the high degree of cross correlation associated with lipidomic data we decided to reduce the dimensionality of the data using Bayesian clustering which has previously been used for lipid datasets. This technique clusters the lipids which behave in a similar fashion together. As expected the lipids largely clustered based on their class. When plotting the average lipid concentration for each cluster only a few clusters showed a change over time. These clusters were: cluster 3 (p = 0.0445) and cluster 12 (p = 0.0089). To see the lipids which are contained in each cluster see Table 2. Lipid cluster 3 contained a broad species of lipids but primarily it contained lipids which were related to inflammation such as Cer, SM and oxidized lipids. Lipid cluster 12 contained large unsaturated TAGs which is a result of the diet. When looking at the association of the lipids and cytokines and clinical characteristics there was a high degree of correlation between the lipid clusters as expected (Figure 5 A&B). At baseline, there were positive correlations between lipid cluster 4 and 6 with IL-6 and CCL3. Lipid cluster 4 contained ceramides and sphingomyelins as well as some diacylglycerols (DAGs), TAGs and unknown TAG species. Lipid cluster 6 contains predominantly unknown TAG species and TAGs. Additionally, there was a positive correlation with lipid cluster 5 and CCL4. There were further positive correlations between IL-1β and lipid cluster 11 and cluster 1 with TNFα. There was also a negative association between TNFα and lipid clusters 4, 5 and 8. Furthermore, there was a negative association between lipid cluster 1 and IL-6. At baseline the only lipid cluster which positively correlated to the clinical features was lipid cluster 4. This lipid cluster was associated with a history of epilepsy prior to admission for SRSE. For more information about the other lipid classes see Table 2. After 1 week of the ketogenic diet there was an even stronger correlation between the lipid clusters suggesting that the diet may alter lipid metabolism in patients with SRSE. Interestingly, the patterns observed with the cytokines changed following diet initiation. Lipid cluster 2 is became positively associated with Il-1β and TNFα. Lipid cluster 2 contains lyso-lipids which have been shown to be altered in a pro-inflammatory state^42^. The only other significant associations between lipids and cytokines were all positive and linked lipid cluster 4 with IL6 and CCL3. CCL3 was also associated with lipid cluster 7, 8, 9, and 10 which contain the TAG species.

**Table 2.**
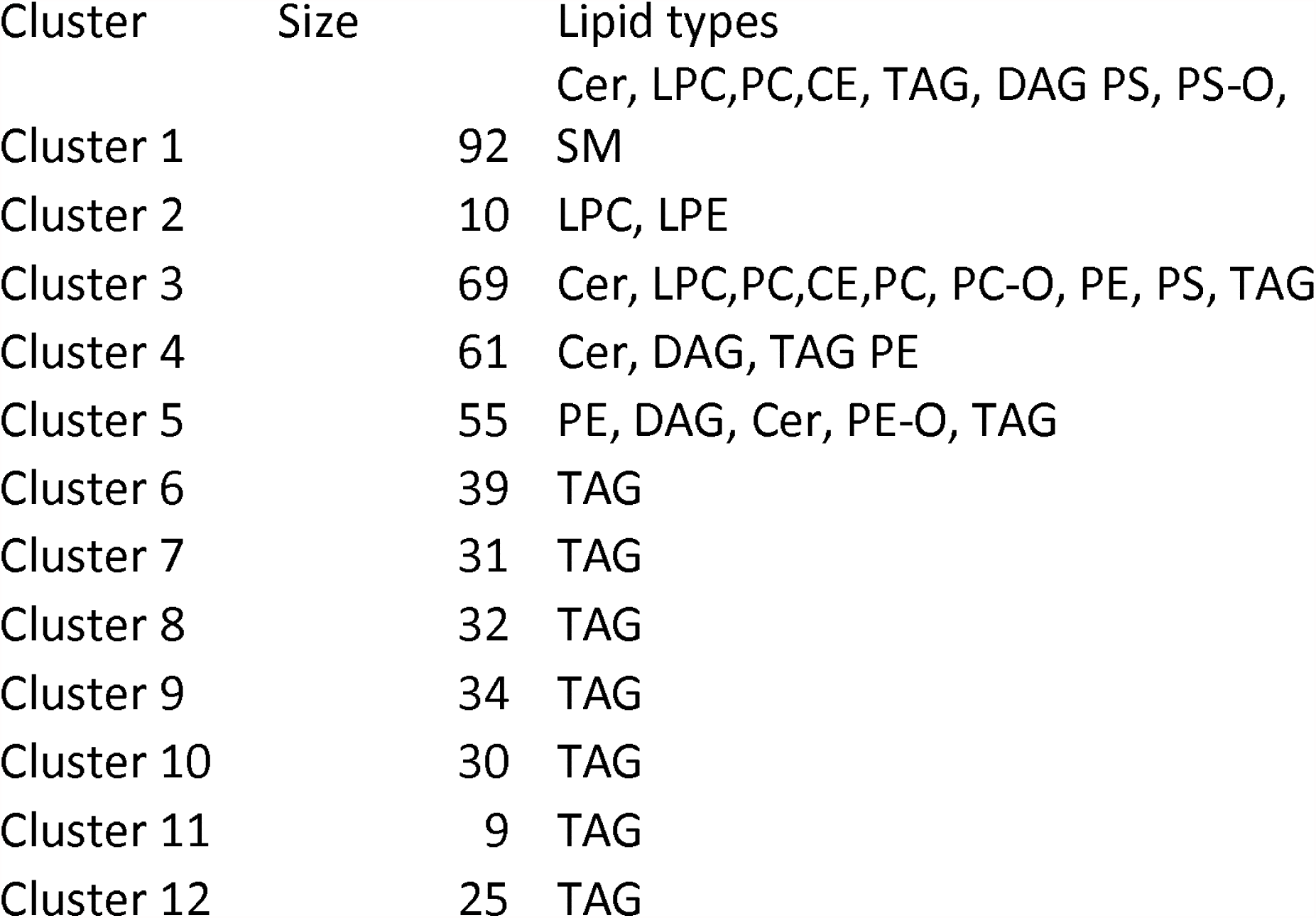

**Figure 5.**
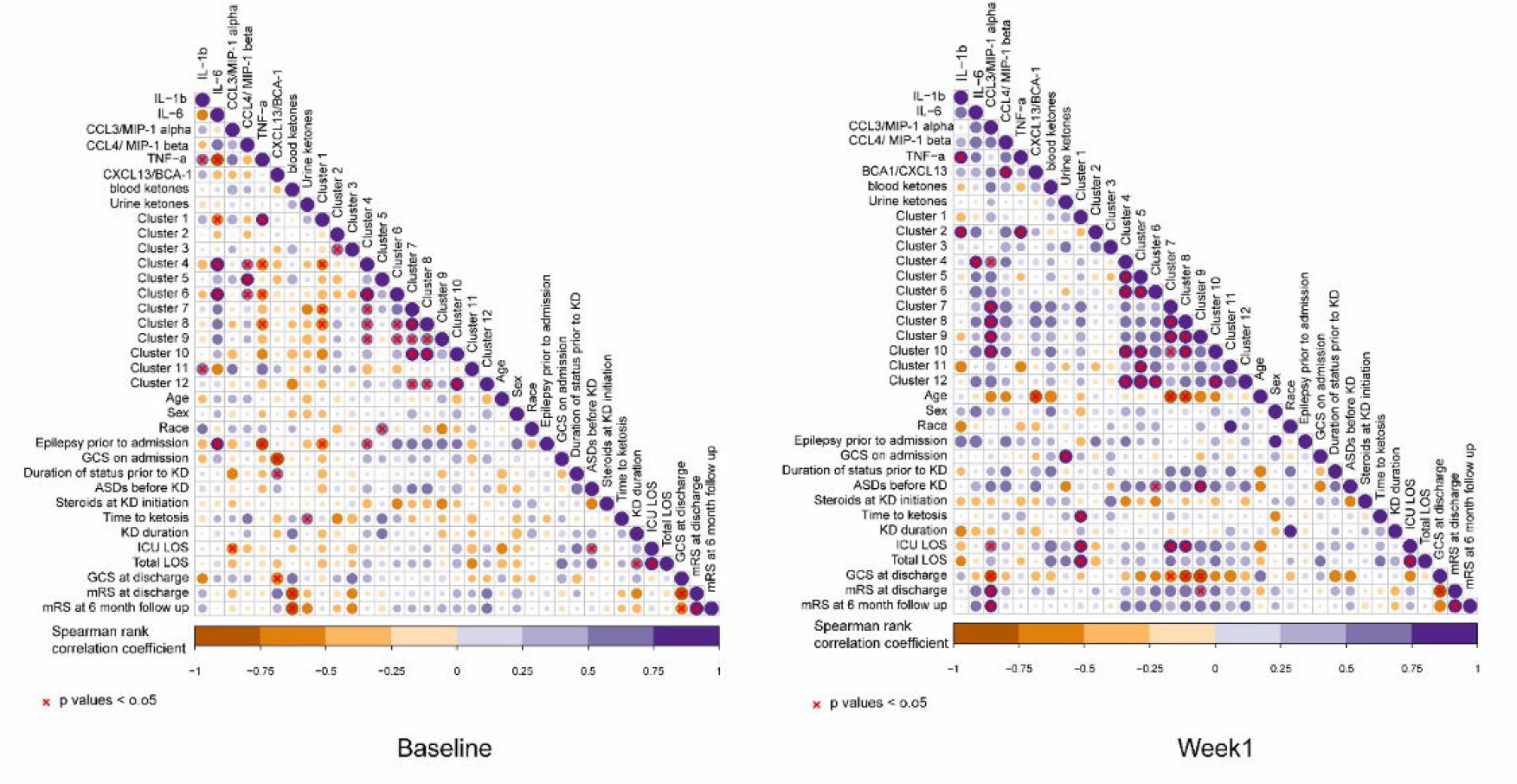
Correlation coefficients illustrated by a heat map. The correlation analysis was performed between lipid clusters and other measurements including cytokine measurements, ketones, patient demographics as well as outcome measure as listed in Fig.1. a) Baseline and b) Week 1 samples. The circles in the plot represents the spearman correlation coefficient between the measurements. Here, purple gradient represents the strength positive correlation while the dark orange gradient indicates strength of negative correlation (red X denote the p values <0.05). The size of circle represent the strength of the correlation coefficient. Bigger circle mean higher correlation coefficient.

### Lipids and inflammatory biomarkers as predictors of clinical outcome

Due to the low number of participants, making predictions on the whole circulating lipidome is very difficult. However, it was possible to build a PLS-DA model using the week 1 blood samples which could differentiate between the participants who recovered with only a mild disability (mRS < 3) compared to those who were moderately or more severely disabled or dead (mRS ≥ 3) (Figure 6A). The cross-validated model had an ROC = 0.56 which is understandable given the very small sample size (n=4 per group). However, from the PLS-DA model it was possible to identify which lipids cause the separation observed in the model by examining the variable importance scores which provides a metric on how each lipid contributes to the PLS-DA model.

**Figure 6.**
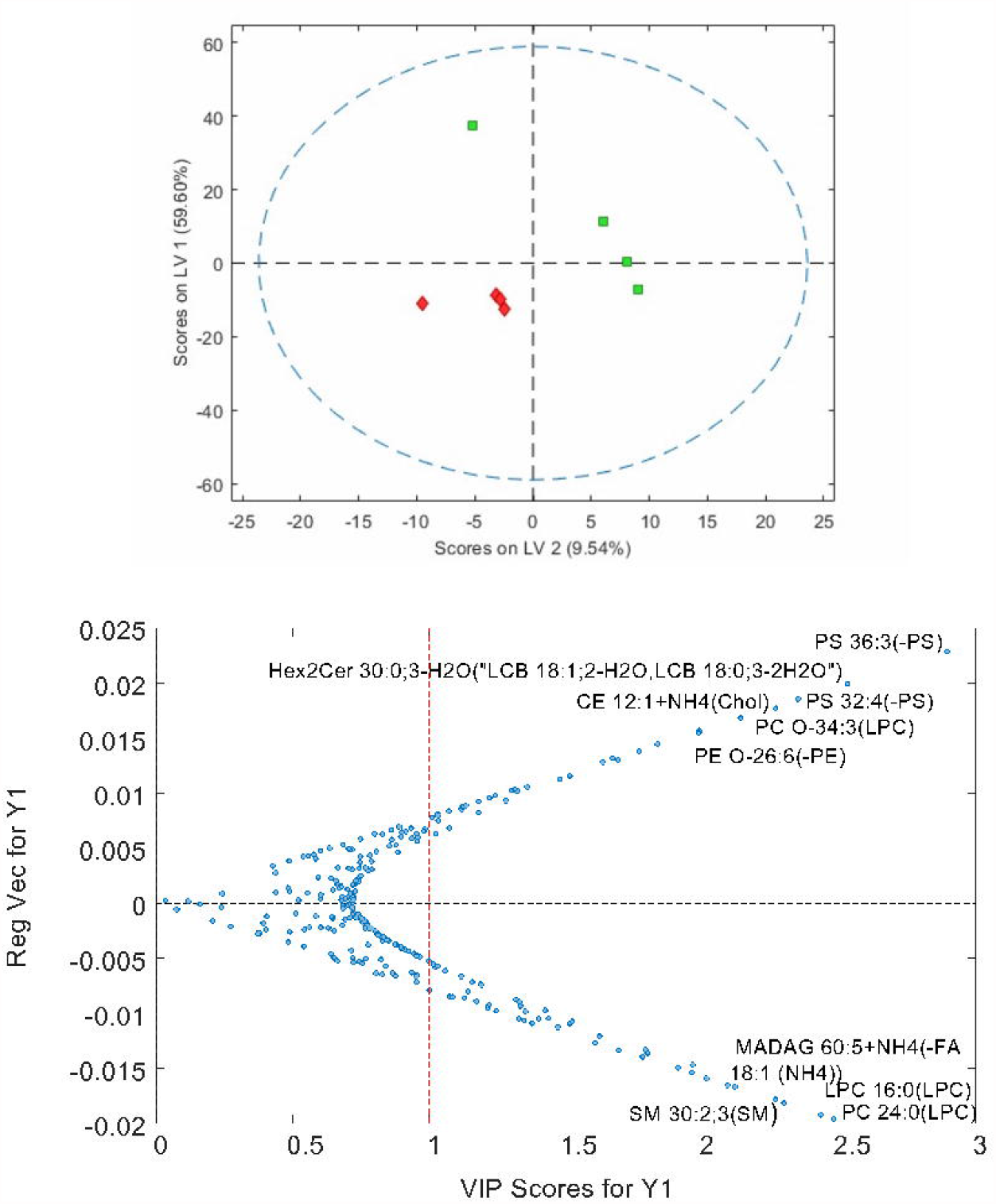
A. PLS-DA model showing the separation of patients who have a good outcome (mRS ≤3, green) compared to patients with a worse outcome (mRS≥4, red). B. Plot showing the variable importance versus the regression vector for the lipids which impact the PLS-DA model. Metabolites at the top are increased in patients who do poorly compared to the lipids at the bottom which are decreased in patients who do poorly.

At baseline (Figure 5), there were very few significant correlations between pro-inflammatory and lipid biomarkers and clinical outcomes. There was a significant negative correlation between GCS score at discharge and CXCL13 levels prior to diet intervention. In other words, those participants with higher CXCL13 levels prior to diet intervention had poorer neurologic outcomes. There was a strong negative correlation between blood β-hyrdroxybutyrate levels at baseline and mRS at discharge and 6-month follow up. In other words, participants with lower blood ketone levels prior to intervention had more severe disability or higher likelihood of death at discharge and at 6 months.

At 1 week, there were several significant correlations between lipid clusters, CCL3 concentrations, and clinical outcomes. While it was previously noted that CCL3 levels positively correlated with lipid clusters 4, 7, 8, 9, and 10 concentrations, higher CCL3 levels also correlated with greater length of ICU stay and higher mRS score (worse neurologic disability) at discharge and 6-month follow up. There was also a significant positive correlation between mRS at discharge and lipid cluster 9. Finally, there was a negative correlation between CCL3 concentrations at 1 week as well as lipid clusters 7, 8, and 9 and GCS scores at discharge.

## Discussion

Super-refractory status epilepticus is a life-threatening disease with no proven treatments. While the ketogenic diet (KD) has been used for nearly one century to manage seizure disorders, it has only been incorporated into the management of refractory and super-refractory status epilepticus in the last decade. This study was designed to determine whether treatment with a ketogenic diet and resultant production of ketone bodies played a role in lipid modulation and reduction in systemic inflammation as potential mechanisms of action for stopping seizures in adults with super-refractory status epilepticus.

Prior to initiation of the ketogenic diet in adults with SRSE, there were several positive correlations between clusters of lipid species and pro-inflammatory cytokine and chemokine levels. There was a specific association with prior history of epilepsy and a lipid cluster which contained ceramides and sphingomyelins as well as some diacylglycerols, triacylglycerols, and monoalkyldiacylglycerol (unknown TAG species), suggesting that the prior epilepsy could have long term effects on these lipids or that these changes could be the underlying cause^43^. The associations of these lipid clusters to pro-inflammatory cytokines^44^ suggests that they may be markers of the underlying inflammation.

Ceramides are a subclass of sphingolipids that are known to regulate the surface expression of NMDA receptors^44^, and to mediate programmed cell death. Ceramides have been shown to increase in hippocampal tissue of adult rats after kainic acid-induced status epilepticus^43, 45^, suggesting a potential role in neuronal injury during status epilepticus. In the study, the levels of ceramides and hexyl ceramides increased over time.

The only biomarker that significantly predicted clinical outcome prior to KD initiation was β-hydroxybutyrate level. Specifically, higher β-hydroxybutyrate levels prior to intervention predicted lower degree of disability at discharge and 6-month follow up. In a lipopolysaccharide-induced fever rodent model, researchers observed a less fever and lower pro-inflammatory cytokines IL-1β and TNFα in animals pre-treated with KD compared to controls although ketone body measures were not included^38^. The authors concluded that KD may have a neuroprotective effect. In RSE and SRSE, it is well-established that early aggressive intervention reduces morbidity and mortality and therefore, participants who were producing ketone bodies prior to KD (either with fasting or other lipid sources pre-diet) may have derived benefit even prior to diet initiation.

There was a significant reduction in systemic pro-inflammatory cytokine IL-6 and pro-inflammatory chemokine CXCL13 on KD. Recent studies have shown that the ketone body β-hydroxybutyrate directly inhibits NLRP3 inflammasome-mediated production of IL-1β^46-48^, reducing systemic inflammation in rodents and human monocytes, which may provide an explanation for the reduction in systemic pro-inflammatory cytokines and chemokines observed in this study. However, urine acetoacetate and blood β-hydroxybutyrate as well as time to ketosis did not show any direct correlation with clinical outcomes or inflammatory biomarker concentrations in this study. Prior studies examining the relationship between ketone body production and seizure cessation in humans have had had mixed results, likely due to a small number of participants^49^.

At the conclusion of the study, higher CXCL13 levels correlated with poorer neurologic outcomes. Recent studies have demonstrated increase in CXCL13 in rodent models of status epilepticus and postulated that this may play a role in pathogenesis^50^. Lower levels of CCL3 and lipid clusters 7-9 predicted better clinical outcome at 1 week of intervention with KD.

This study had several important limitations. The number of participants was small and therefore few significant correlations could be identified. In addition, serial sample collection was not feasible for all participants and IL-1β and TNFα measures were below the detectable range in most participants. Expected changes in lipids and inflammatory cytokine and chemokine concentrations with progression of SRSE are unknown and therefore which changes may have been a result of KD or ketone body production cannot be determined.

In conclusion, this study provides preliminary evidence that lipid modulation and reduction in pro-inflammatory biomarkers occur during administration of a ketogenic diet in adults with super-refractory status epilepticus. Furthermore, lower levels of pro-inflammatory chemokines and certain lipid species predict better neurologic outcome following KD. Further studies are needed to understand the potential neuroprotective effects of lipid modulation an reduction in inflammation using KD in the setting of refractory status epilepticus and to establish the relationship between modulation of specific lipid species, inflammation, ketone body production, and seizure cessation.

## Supporting information

STROBE Checklist

## Data Availability

The data is avlible upon request

## Acknowledgements

We would like to thank the patients and families who participated in this study. This study was funded by the generous philanthropic support through the Johns Hopkins Center for Status Epilepticus and Neuroinflammation from Chris Garrod and Dawn Griffiths. We would also like to thank Jacqueline Lovett for sample collection and processing and Bareng Nonyane and Ximin Li for biostatistical support.

## Study Funding

This study was funded by the Johns Hopkins Center for Status Epilepticus and Neuroinflammation.

## Disclosures

1. Alex Dickens, PhD

Disclosure: Dr Dickens has worked part time for Orion Pharma (Finland). This study has nothing to do with his work at this company.

2. Tory Johnson, MD

Disclosure: Dr. Johnson has no relevant disclosures.

3. Santosh Lamichhane, PhD

Disclosure: Dr. Lamichhane has no relevant disclosures.

4. Anupama Kumar, MBBS

Disclosure: Dr. Kumar has no relevant disclosures.

5. Carlos A. Pardo, MD

Disclosure: Dr. Pardo has no relevant disclosures.

6. Erie G. Gutierrez, MD

Disclosure: Dr. Gutierrez has no relevant disclosures.

7. Norman Haughey, PhD

Disclosure: Dr. Haughey has no relevant disclosures.

8. Mackenzie C. Cervenka, MD

Disclosure: Dr. Cervenka receives grants from Nutricia, Vitaflo, BrightFocus Foundation, and Army Research Laboratory. Honoraria from American Epilepsy Society, The Neurology Center, Epigenix, LivaNova, and Nutricia. Royalties from Demos Health/Springer Publishing Company. Consulting for Nutricia, Sage Therapeutics.

## Notes

### Competing Interest Statement

Alex Dickens, DPhil
Disclosure: Dr Dickens has worked part time for Orion Pharma (Finland). This study has nothing to do with his work at this company.
Tory Johnson, PhD
Disclosure: Dr. Johnson has nothing relevant to disclose.
Mackenzie C. Cervenka, MD
Disclosure: Dr. Cervenka receives or has received grants from Nutricia, Vitaflo, BrightFocus Foundation, and Army Research Laboratory. Honoraria from American Epilepsy Society, The Neurology Center, Epigenix, LivaNova, and Nutricia. Royalties from Demos/Springer. Consulting for Nutricia, Sage Therapeutics.

### Author Declarations

Informed consent was obtained from a legally authorized representative and the Johns Hopkins Medicine Institutional Review Board approved the study (IRB # NA_00073063 and NA_00003551).

